# A Multicomponent Metabolic Formulation in Adults With Major Depressive Disorder: An Exploratory Interim Analysis of a Randomized Mechanistic Trial

**DOI:** 10.64898/2026.11.25.26368337

**Authors:** Martin P Paulus, Rayus Kuplicki, Hung-Wen Yeh, Elena C Gross

## Abstract

Metabolic interventions are being investigated for major depressive disorder (MDD), but evidence for specific formulations remains limited. This double-masked, placebo-controlled randomized trial evaluates a multicomponent formulation containing D-β-hydroxybutyrate (D-BHB), micronutrients, and metabolic cofactors for 8 weeks. The registered primary outcome is week 8 nucleus accumbens blood-oxygen-level-dependent response during Monetary Incentive Delay reward anticipation; the secondary outcome is medial prefrontal response during self-referential processing. Here, we report an exploratory Bayesian interim analysis of 62 individuals screened, 29 randomized (active, n=15; placebo, n=14), 26 who received at least one dose (safety population), and 22 who were evaluable (active, n=10; placebo, n=12). D-BHB rose after week 8 dosing in the active arm but not placebo. The primary week 8 active-minus-placebo estimate was 0.02 (95% credible interval [CrI], −0.12 to 0.17); the difference in change was 0.00 (−0.17 to 0.18). The covariate-adjusted secondary estimates were 0.03 (−0.13 to 0.18) at week 8 and 0.00 (−0.18 to 0.17) for change. Exploratory clinician-rated symptoms favored active treatment (MADRS week 8 difference, −7.28 [−12.00 to −2.47]), whereas self-reported symptoms were uncertain (QIDS-SR16, −1.95 [−5.13 to 1.15]). Three of 26 exposed participants reported four mild adverse events; none was serious or judged treatment-related. The formulation increased circulating D-BHB, but engagement of the prespecified imaging targets was not demonstrated. The exploratory clinician-rated symptom signal requires confirmation and cannot be attributed to D-BHB alone.

## Introduction

Major depressive disorder (MDD) remains a major cause of disability, and treatment response using standard antidepressant interventions is often incomplete [1, 2].

MDD has increasingly been associated with disturbances in cellular bioenergetics, mitochondrial function, oxidative stress, and inflammatory regulation, suggesting that impaired energy metabolism may represent a therapeutically relevant mechanism in at least a subset of patients [3–5]. Mitochondrial abnormalities reported in MDD include alterations in energy production and redox homeostasis and interactions between mitochondrial dysfunction and inflammatory signaling, although their causal contribution and clinical relevance remain incompletely understood [3–5]. Cellular energy production depends among other factors on an interconnected network of vitamins, minerals, and mitochondrial cofactors required for substrate oxidation, electron transport, ATP generation, redox homeostasis, one-carbon metabolism, and neurotransmitter synthesis [6]. Several metabolically relevant nutrients and cofactors have therefore been investigated in depression, with randomized trials and meta-analyses reporting signals of benefit for interventions including L-methylfolate, magnesium, zinc, vitamin D, and coenzyme Q10, although findings vary by nutrient, population, and study design [7–12]. Together, these observations provide a rationale for investigating metabolic interventions that address both energy-substrate availability and the micronutrient/cofactor requirements of cellular energy metabolism.

Nutritional ketosis and exogenous ketones have been proposed as lower-burden metabolic interventions. D-β-hydroxybutyrate (D-BHB) can serve as an energy substrate and signaling metabolite; preclinical work implicates inflammasome, redox, epigenetic, transport, and neurotrophic pathways [13–17]. Early clinical studies of ketogenic dietary interventions in depression are encouraging but remain limited [18, 19]. Moreover, it is not clear whether the exposure achieved with an exogenous ketone formulation has similar effects as sustained nutritional ketosis.

The formulation tested here was developed according to this multicomponent metabolic approach. Rather than providing D-BHB in isolation, it combines D-BHB as an alternative oxidative energy substrate with approximately 30 additional micronutrients and metabolic/mitochondrial cofactors. The rationale for the formulation is therefore to address both energy-substrate availability and the metabolic machinery supporting cellular energy metabolism. The present trial evaluates the complete multicomponent metabolic formulation rather than D-BHB as an isolated intervention.

This randomized mechanistic trial examines whether an oral multicomponent metabolic formulation containing D-BHB, micronutrients, and metabolic cofactors engages two task-fMRI targets in adults with MDD enriched for low-grade inflammation. The registered primary outcome is nucleus accumbens (NAcc) activation during reward anticipation; the registered secondary outcome is medial prefrontal cortex (mPFC) activation during self-referential processing. We report an exploratory Bayesian interim analysis. The analysis was undertaken for preliminary reporting without a prespecified stopping rule; the trial remains ongoing.

## Methods

### Design, setting, registration, and oversight

This investigator-initiated, single-site, parallel-group randomized trial was conducted at the Laureate Institute for Brain Research (Tulsa, Oklahoma). Recruitment began January 9, 2024. Participants were assigned 1:1 to active supplement or matched placebo for 8 weeks. The trial was registered as NCT06263660; the current public record was accessed August 28, 2026 [20]. Protocol version was dated October 30, 2023. No separate statistical analysis plan (SAP) was supplied. The institutional review board approved the protocol (WCG IRB Protocol #20225936), and all participants provided written informed consent. The trial remains ongoing; the registry currently lists estimated enrollment of 75 to obtain 60 completers, estimated primary completion December 31, 2026, and estimated study completion December 31, 2027.

### Participants

Eligible participants were adults aged 18–65 years with current nonpsychotic MDD confirmed by the Mini International Neuropsychiatric Interview, Patient Health Questionnaire-9 score ≥10, C-reactive protein >1 mg/L, body mass index 19–38 kg/m^2^, and stable psychotropic medication dose and frequency for 6 weeks before entry. Key exclusions included clinically significant suicidality, current psychotic or bipolar disorder, recent moderate-to-severe substance use disorder, MRI contraindication or inability to complete MRI, pregnancy or breastfeeding, and medical instability.

### Randomization and masking

Block randomization with block size 10 was used. Product was labeled Arm A or Arm B. Participants, investigators, recruitment personnel, clinicians, and outcome assessors were intended to remain masked; designated unmasked study staff maintained allocation and dispensed product. Analysts worked with A/B labels. Post-dose D-BHB patterns later identified Arm A as active, producing de facto unmasking from the biochemical data. Participant guesses and certainty were collected; a descriptive arm difference in high certainty (Fisher exact P=0.12) raises, but does not establish, a masking concern.

### Interventions

The active intervention was the KetoSwiss Brain Energy Complex formulation, a multicomponent metabolic supplement developed and supplied by KetoSwiss AG, with a nominal content of 5 g D-BHB as calcium and magnesium salts per approximately 9.25-g packet, together with micronutrients and metabolic cofactors. The reported composition and analytical testing results for lot 23214A (9.2-g analytical serving) are presented in Supplementary Tables S16 and S17. Placebo was dextrose-based and matched for appearance and administration. Participants dissolved one packet in water three times daily for 8 weeks. At baseline and week 8 imaging visits, two packets were taken together before scanning and the third dose was taken that evening.

### Outcomes

The registered primary outcome was NAcc blood-oxygen-level-dependent (BOLD) activation during reward anticipation on the Monetary Incentive Delay (MID) task, operationalized as P5 versus P0 in bilateral Brainnetome regions 223 and 224. The registered secondary outcome was mPFC BOLD activation during self-referential processing, operationalized as SELF versus WORD in Brainnetome regions 13–14, 41–42, and 187–188. Both were measured at baseline before dosing (V0Pre), baseline after dosing (V0Post), and week 8 (V4). The week 8 between-arm contrast was specified for both primary and secondary outcomes. MADRS and QIDS-SR16 symptom scales, adherence, biochemical exposure, masking assessments, and adverse events were treated as exploratory, feasibility, or safety measures in this report. Follow-up analyses on primary, secondary, and other endpoints included the treatment by time (V4 – V0Pre) interaction.

### Neuroimaging acquisition and processing

Images were acquired on a Siemens 3-T Prisma scanner with a 20-channel head coil. Functional echo-planar imaging used repetition time 1500 ms, echo time 27 ms, flip angle 72°, 2.4-mm isotropic voxels, 54 slices, and multiband factor 2. MID comprised two 375-volume runs; the adjective task comprised 476 volumes. Processing used AFNI 24.3.10 [21]: slice-time and rigid-motion correction, skull stripping, rigid T1–EPI registration, 12-parameter affine normalization to MNI space, and 4-mm full-width-at-half-maximum smoothing. Volumes were censored when ENORM exceeded 0.3 mm or outlier fraction exceeded 0.1; runs were discarded when mean ENORM exceeded 0.3 mm. First-level models included motion and fourth-order Legendre nuisance terms. Regions were defined independently with Brainnetome Atlas v1.0 [22]. Additional acquisition, task, and QC details appear in Supplementary Methods.

### Adherence, biochemical exposure, and safety

Packet-count adherence was 100 × (packets dispensed − packets returned) / {3 × [V4 date − V0 date] − 2}. The −2 accounts for two baseline packets administered in clinic rather than taken from dispensed supplies. The actual interval was 52–63 days. One value was 100.6%; it was verified against accountability records and analyzed after capping at 100%, with exclusion as sensitivity analysis. Self-report adherence and electronic-cap metrics specified elsewhere were not used in the interim analysis.

Week 8 D-BHB was measured before dosing and 20, 40, and 80 minutes after dosing. The assay lower limit of quantification (LLOQ) was 0.05 mmol/L and we imputed 0.05/√2 for below-LLOQ values. No biochemical exposure threshold was prospectively defined. The safety population included all randomized participants receiving ≥1 dose. Adverse events through the December 19, 2025 freeze were summarized descriptively from free text; no MedDRA coding was applied in the interim dataset.

### Analysis populations and exploratory interim-analysis status

The randomized population comprised 29 participants. Three active-arm participants never received product; the safety population therefore comprised 26 exposed participants. Four exposed participants did not enter the evaluable set because of inability to complete intervention/imaging (n=1), scheduling (n=2), or inability to complete a blood draw (n=1). The evaluable and primary-analysis population comprised 22 participants (active, n=10; placebo, n=12). Primary-outcome data were complete at all three visits; secondary-outcome data were available for 22 at V0Pre, 20 at V0Post, and 19 at week 8.

The protocol specified an interim analysis after approximately 10 participants in the per-protocol dataset, without efficacy, futility, safety, or target-engagement boundaries and without use to determine continuation using a non-Bayesian approach. The Bayesian approach described below and current 22-person dataset therefore cannot be established as prospectively specified. We describe this report as an unplanned exploratory Bayesian interim analysis; registered outcomes and measurement times remain prespecified design elements.

### Bayesian statistical analysis

All time-dependent outcomes were analyzed by Gaussian identity-link mixed models including arm, visit, and arm-by-visit interaction as fixed effects and participant as a random intercept. Repeated observations were modeled using all available data under a missing-at-random assumption. Contrasts were active minus placebo at week 8 and the between-arm difference in change from V0Pre to week 8. Posterior medians and equal-tailed 95% credible intervals (CrIs) are reported; posterior probabilities are descriptive, and no decision threshold or multiplicity adjustment was defined. The secondary-outcome model adjusted for age, sex, body mass index, and medication use. MADRS and QIDS-SR16 estimates shown here are unadjusted exploratory models.

Adherence was modeled in brms using a one-inflated beta distribution constrained to allow proportion 1 (100%) adherence; the mean and boundary components included arm. All Bayesian models used four chains, 4000 iterations per chain, including 2000 warm-up iterations, no thinning, and 2000 post-warm-up draws per chain. Modeling used R 4.5.0, rstanarm 2.32.1, brms 2.23.0, and marginaleffects 0.28.0.

Reported maximum R-hat values were 1.004 or lower, minimum bulk effective sample size was 1442, and no divergent transitions or maximum treedepth events were reported. Posterior predictive checks were broadly compatible with observed distributions.

## Results

### Participant disposition and baseline characteristics

Sixty-two people were assessed, 33 were excluded, and 29 were randomized (active, n=15; placebo, n=14). Twenty-six received at least one dose (active, n=12; placebo, n=14), and 22 were evaluable (active, n=10; placebo, n=12) (Fig. 1). Median age in the evaluable set was 41 years (interquartile range [IQR], 27–49), 18 (82%) were female, and 12 (55%) used psychotropic medication (Table 1). Baseline active-arm imaging data had more censored repetition times than placebo for both tasks; this preceded exposure and was interpreted as chance imbalance. No participant was excluded for an extreme endpoint value.

**Figure 1.**
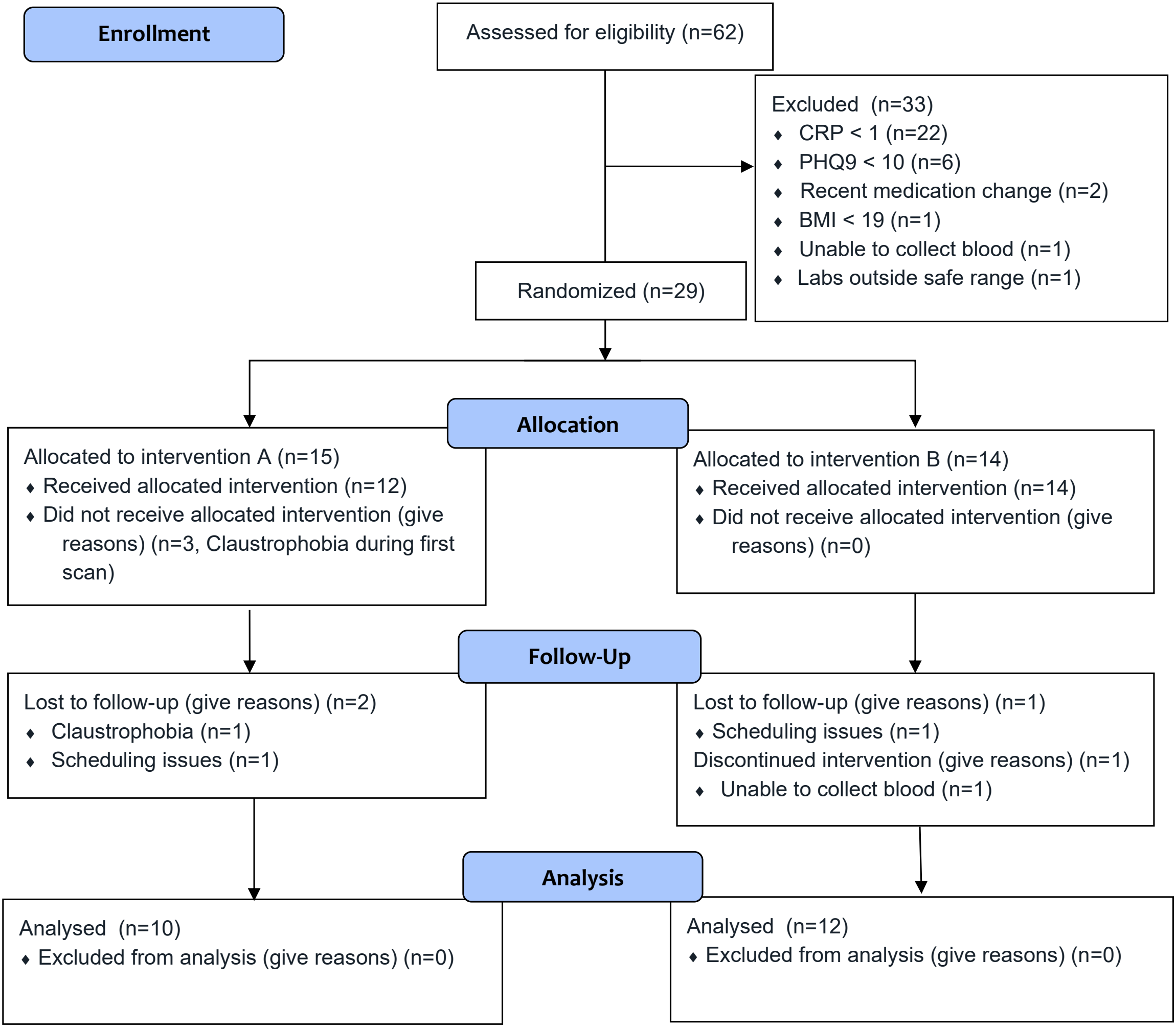
Participant flow through the interim data freeze. The randomized population was 29, the safety population included 26 participants receiving at least one dose, and the evaluable/primary analysis population included 22. Active and placebo assignments were inferred from post-dose D-BHB patterns after analyses had been conducted under A/B labels.

**Table 1.** Baseline characteristics of the evaluable interim sample.

| **Characteristic** | **Overall (N=22)** | **Active (n=10)** | **Placebo (n=12)** |
|---|---|---|---|
| Age, median (IQR), y | 41 (27–49) | 40 (25–54) | 41 (33–48) |
| Female, n (%) | 18 (82) | 8 (80) | 10 (83) |
| Male, n (%) | 4 (18) | 2 (20) | 2 (17) |
| BMI, median (IQR), kg/m<sup>2</sup> | 32.2 (26.1–35.0) | 30.3 (22.3–34.9) | 33.5 (31.0–35.5) |
| Psychotropic medication use, n (%) | 12 (55) | 5 (50) | 7 (58) |
| MADRS, median (IQR) | 21.0 (18.0–25.0) | 20.0 (18.0–26.0) | 21.5 (19.0–24.5) |
| QIDS-SR16, median (IQR) | 11.0 (8.0–14.0) | 11.5 (8.0–15.0) | 11.0 (9.5–13.0) |

BMI, body mass index; IQR, interquartile range; MADRS, Montgomery–Åsberg Depression Rating Scale; QIDS-SR16, Quick Inventory of Depressive Symptomatology Self-Report. No baseline hypothesis tests are presented.

### Exposure and adherence

Week 8 D-BHB data were available for 21 of 22 evaluable participants. Active-arm medians rose from <0.05 mmol/L before dosing to 0.15 at 20 minutes, 0.16 at 40 minutes, and 0.09 at 80 minutes; corresponding placebo medians were <0.05, <0.05, 0.05, and <0.05 mmol/L. Median peak post-dose D-BHB was 0.18 mmol/L (range, 0.08–0.28) with active treatment and <0.05 (<0.05–0.25) with placebo (Figure S1). These data identify biochemical separation but cannot establish adequacy because no target concentration was specified.

Packet-count adherence was high: active mean 90.1% (SD 13.0; median 97.2%; range 65.6–100.6) and placebo mean 91.9% (SD 7.2; median 92.8%; range 78.9–100.0). In the capped one-inflated beta model, the active-minus-placebo average marginal effect was 0.012 (95% CrI, −0.071 to 0.094); excluding the 100.6% value gave 0.001 (−0.086 to 0.084).

### Primary and secondary imaging outcomes

The primary NAcc MID endpoint did not separate the groups. The active-minus-placebo estimate at week 8 was 0.02 (95% CrI, −0.12 to 0.17), and the difference in change was 0.00 (−0.17 to 0.18); posterior probabilities that the effects exceeded zero were 0.630 and 0.517, respectively (Table 2; Fig. 2).

**Table 2.** Bayesian estimates.

| Outcome | Estimand | Active − placebo, median (95% CrI) | Posterior probability | Model / data |
|---|---|---|---|---|
| NAcc MID (primary) | Week 8 | 0.02 (−0.12 to 0.17) | 0.630 | Unadjusted; N=22, 66 observations |
| NAcc MID (primary) | Change | 0.00 (−0.17 to 0.18) | 0.517 | Unadjusted; N=22, 66 observations |
| mPFC SELF–WORD (secondary) | Week 8 | 0.00 (−0.18 to 0.18) | 0.526 | Unadjusted*; N=22, 61 observations |
| mPFC SELF–WORD (secondary) | Change | 0.08 (−0.15 to 0.33) | 0.745 | Unadjusted*; N=22, 61 observations |
| MADRS (exploratory) | Week 8 | −7.28 (−12.00 to −2.47) | 0.998† | Unadjusted; N=22 |
| MADRS (exploratory) | Change | −7.12 (−12.80 to −1.52) | 0.993† | Unadjusted; N=22 |
| QIDS-SR16 (exploratory) | Week 8 | −1.95 (−5.13 to 1.15) | 0.886† | Unadjusted; N=22 |
| QIDS-SR16 (exploratory) | Change | −1.88 (−4.88 to 1.13) | 0.897† | Unadjusted; N=22 |
| Adherence (feasibility) | Average marginal effect | 0.012 (−0.071 to 0.094) | — | Capped at 100%; N=22 |

BOLD outcomes are in extracted contrast units; symptom outcomes are scale points; adherence is a proportion.
\*Adjusted for age, sex, BMI, and medication use because an unadjusted July 29 secondary model was not supplied.
†Probability that active treatment produces a lower symptom score. CrI, credible interval; MID, Monetary Incentive Delay; mPFC, medial prefrontal cortex; NAcc, nucleus accumbens.

**Figure 2.**
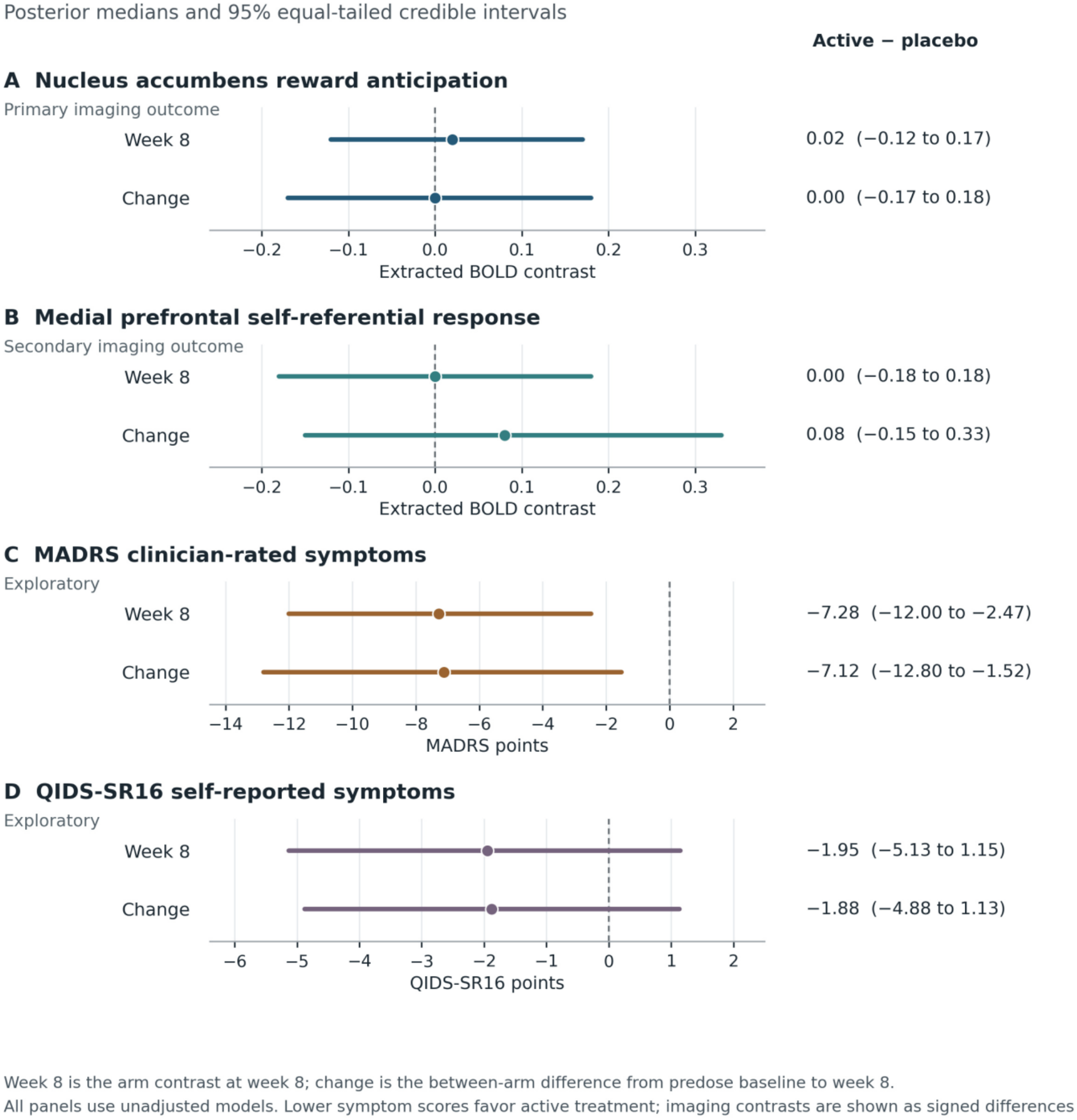
Posterior estimates. Points are posterior medians and lines are equal-tailed 95% credible intervals for active minus placebo. Week 8 is the cross-sectional arm contrast; change is the arm difference in change from V0Pre to week 8. The secondary mPFC model is adjusted for age, sex, BMI, and medication use. Lower symptom scores favor active treatment. MID, Monetary Incentive Delay; mPFC, medial prefrontal cortex; NAcc, nucleus accumbens; MADRS, Montgomery–Åsberg Depression Rating Scale; QIDS-SR16, Quick Inventory of Depressive Symptomatology Self-Report.

For the secondary mPFC self-referential endpoint, the active-minus-placebo estimates were 0.00 (95% CrI, −0.18 to 0.18) at week 8 and 0.08 (−0.15 to 0.33) for difference in change. These intervals include effects in both directions and do not support target engagement.

### Exploratory symptoms

MADRS favored active treatment in the exploratory unadjusted model: the week 8 difference was −7.28 points (95% CrI, −12.00 to −2.47; posterior probability of a lower active score, 0.998), and the difference in change was −7.12 (−12.80 to −1.52; probability, 0.993). QIDS-SR16 estimates were smaller and uncertain: −1.95 (−5.13 to 1.15) at week 8 and −1.88 (−4.88 to 1.13) for change. Adjustment for participant drug-certainty ratings attenuated the MADRS association; because certainty was measured after randomization and may reflect treatment experience, that analysis changes the estimand and is a diagnostic rather than a confounder-adjusted causal analysis.

### Safety

Three of 26 exposed participants reported four adverse events through the data freeze (Table 3). All were mild; none was serious, severe, or judged related to study product, and none led to interruption, discontinuation, withdrawal, or death. Three participants reported passive or transient suicidal ideation without behavior, intent, or plan (active, 1/12; placebo, 2/14). One placebo participant felt unwell during MRI; the event resolved after food and rest.

**Table 3.** Adverse events (safety population)

| **Category** | **Active (n=12)** | **Placebo (n=14)** | **Total (N=26)** |
|---|---|---|---|
| Participants with any adverse event | 1 (8.3) | 2 (14.3) | 3 (11.5) |
| Total adverse events, number | 1 | 3 | 4 |
| Treatment-related adverse events | 0 | 0 | 0 |
| Serious or severe adverse events | 0 | 0 | 0 |
| Dose interruption/discontinuation/withdrawal | 0 | 0 | 0 |
| Deaths | 0 | 0 | 0 |
| Suicidal ideation or behavior | 1 (8.3) | 2 (14.3) | 3 (11.5) |
| Constitutional event (feeling unwell) | 0 | 1 (7.1) | 1 (3.8) |

Values are participants, n (%), unless stated. Percentages use exposed participants in each arm. Events were recorded as free text and were not MedDRA coded in the interim dataset. All reported events were mild and assessed as not related or unlikely related.

## Discussion

This exploratory interim analysis demonstrated a modest post-dose rise in circulating D-BHB but did not detect engagement of either prespecified task-fMRI target. Exploratory clinician-rated symptoms favored the multicomponent formulation at week 8 (MADRS difference, −7.28 points; 95% CrI, −12.00 to −2.47). The self-reported QIDS-SR16 difference was less certain. This clinical signal warrants further study, but the small interim sample, evaluable-subset analysis, and exploratory outcome status preclude a conclusion of antidepressant efficacy.

A null regional task contrast does not establish absence of all biological activity. D-BHB could influence distributed bioenergetic, inflammatory, mitochondrial, or network processes not captured by the selected regions or tasks [13–17]. On the other hand, evidence of peripheral exposure does not demonstrate brain uptake or a therapeutic mechanism. Moreover, the observed concentrations were modest and transient compared with sustained nutritional ketosis. These findings leave unresolved whether the lack of detectable target engagement reflects insufficient central exposure, limited sensitivity of the selected imaging endpoints, or absence of an effect on the hypothesized neural targets.

The symptom pattern should be considered alongside other ketogenic-diet studies [18, 19], although the current intervention differs importantly in that it consisted of a multicomponent metabolic formulation rather than D-BHB alone. D-BHB represents one mechanistically relevant component, providing an alternative oxidative substrate and acting as a signaling metabolite [13–17]; however, the formulation also contains approximately 30 active micronutrients and metabolic cofactors selected to support cellular and mitochondrial metabolism. Several of these components participate in pathways relevant to cellular bioenergetics, including mitochondrial substrate oxidation, electron transport, ATP-dependent reactions, one-carbon metabolism, and redox regulation [6]. Moreover, several individual nutrients or nutrient classes represented in the formulation have independently shown signals of benefit in randomized studies and meta-analyses of depression, including L-methylfolate [7, 8], magnesium [9], zinc [10], vitamin D [11], and CoQ10 [12]. Thus, the biological activity of the formulation cannot be assumed to derive predominantly from D-BHB. The present study was not designed to determine the contribution of individual ingredients or interactions among them, and the relative contributions of D-BHB, individual micronutrients, and their combination therefore remain unresolved.

The MADRS difference was attenuated in a sensitivity analysis adjusting for perceived treatment certainty. However, because treatment certainty was assessed after randomization and may itself have been influenced by treatment experience, this analysis should be interpreted as a diagnostic sensitivity analysis rather than as conventional adjustment for a baseline confounder. Accordingly, the symptom findings remain exploratory and require confirmation in the completed trial.

This study has several strengths which include randomized allocation, intended masking, independently defined imaging regions, biochemical exposure measurement, complete primary-outcome observations among the evaluable set, explicit distinction between randomized, exposed, and evaluable populations, and detailed imaging quality control. Versioned executable analysis and diagnostic outputs enabled numerical cross-checking.

However, there are also limitations. This is a single-site, small, exploratory interim analysis rather than the final registered trial analysis. Analyses used an evaluable rather than randomized population, creating potential post-randomization selection. The secondary model was covariate-adjusted, missing observations were assumed missing at random without sensitivity analysis, and multiple endpoints and sensitivities were examined without a prespecified decision framework. Biochemical data created de facto unmasking. The intervention is multicomponent, exposure was modest, and the enriched sample limits generalizability.

This exploratory interim analysis did not detect engagement of the prespecified fMRI targets, while clinician-rated symptoms favored the multicomponent metabolic formulation. The clinical signal warrants evaluation in the completed trial and independent replication. Whether any benefit depends on D-BHB, other ingredients, their combination, or expectancy-related effects remains unresolved.

## Data Availability

All data produced in the present study are available upon reasonable request to the authors.

## Acknowledgements

The authors would like to acknowledge Dara Crittenden for her invaluable help in conducting the study.

## Author contributions

Conceptualization: Paulus, Gross. Investigation and data interpretation: all authors. Statistical analysis: Kuplicki, Yeh. Original draft: Paulus. Review and editing: all authors. Funding acquisition and supervision: Paulus.

## Ethics approval

The study was approved by the WCG IRB and all participants provided written informed consent.

