## Supplementary Information for "A Multicomponent Metabolic Formulation in Adults With Major Depressive Disorder: An Exploratory Interim Analysis of a Randomized Mechanistic Trial"

ClinicalTrials.gov: NCT06263660 | Data freeze/database lock: 19 December 2025 | Analysis report: version 909, 29 July 2026

### Contents

- Supplementary Methods: chronology; source hierarchy; design and outcomes; imaging; analysis populations; Bayesian analysis; missing data; adherence; biochemical exposure; masking; safety; formulation composition and analytical testing
- Supplementary Results: diagnostics; sensitivity analyses; imaging quality control; biochemical exposure; adherence; adverse-event listing
- Supplementary Tables S1–S17
- Supplementary Figures S1–S3
- Transparency summary and provenance

### Supplementary Methods

#### S1. Trial and analysis chronology

The chronology below separates documented dates from unresolved or current registry dates. The supplied protocol itself, amendment history, and contemporaneous analytical correspondence were not included, so absence of documentation is not evidence that an event did not occur.

Table S1. Chronology

| **Date** | **Event** | **Source** | **Status / implication** |
| --- | --- | --- | --- |
| 30 Oct 2023 | Protocol version 3.0 dated | Henry/Rayus interim memoranda | Documented; protocol file not supplied |
| 9 Jan 2024 | Study start | Current ClinicalTrials.gov record | Registry date |
| 16 Feb 2024 | First public posting/listing | Registry history mirror/current record metadata | Confirm direct history download; start preceded first posting |
| 19 Dec 2025 | Interim data freeze and database lock | 30 Jul memo; Henry memo | Documented; exact last safety follow-up date unknown |
| 29 Jul 2026 | Version 729 Bayesian report | Executable HTML | Authoritative numerical analysis supplied |
| 30 Jul 2026 | Interim-status memorandum | Rayus memo | Documents freeze, no SAP, de facto unmasking |
| 31 Jul 2026 | Adherence/BHB memorandum | Adherence memo | Corrects adherence estimate and 80-min draw |
| 28 Aug 2026 | Registry and NPP requirements checked | Current public pages | Submission-preparation date |
| 9 Sep 2026 | Version 929 Bayesian report | Henry memo | Updating 2,000 iteration for non-primary outcomes in v729 with 4,000 iterations |
| 31 Dec 2026 | Estimated primary completion | Current registry | Future/current estimate |
| 31 Dec 2027 | Estimated study completion | Current registry | Future/current estimate |

#### S2. Source hierarchy and governing documents

Numerical estimates were taken from the July 29, 2026 version 909 executable output unless a later memo explicitly corrected a descriptive measure. Participant disposition came from the participant-level disposition workbooks and CONSORT source. Adverse-event counts came from the dedicated safety file. Imaging methods and QC came from the dedicated methods/QC memoranda. The July 30 and July 31 memoranda superseded conflicting narrative statements in the May manuscript. ClinicalTrials.gov governed current registration fields.

Table S2. Source hierarchy

| **Domain** | **Controlling source** | **Boundary** |
| --- | --- | --- |
| Trial design and outcome registration | ClinicalTrials.gov current record; protocol references in memos | Protocol and amendments not supplied |
| Analysis estimates | Version 909 executable report (29 Jul 2026) | Rerun needed for exact priors/settings and unadjusted secondary model as well as the consistent 4,000 iterations for all outcomes |
| Disposition/populations | Participant-level Excel files; legacy CONSORT | 29 randomized / 26 exposed / 22 evaluable |
| Adherence and D-BHB | 31 Jul 2026 memorandum | Supersedes older adherence effect |
| Safety | Adverse Events Main Table and eTable | Exact last follow-up and MedDRA coding missing |
| Imaging endpoints/methods/QC | Endpoint, eMethods, and QC memoranda | Endpoint-status wording internally inconsistent |

#### S3. Registered and analyzed outcomes

Table S3. Outcome definitions and status

| **Classification** | **Measure** | **Time** | **Estimand/reporting** | **Status** |
| --- | --- | --- | --- | --- |
| Primary | NAcc MID P5 vs P0 | V0Pre, V0Post, week 8 | Week 8 active−placebo; change contrast also reported | Registered; inferential contrast awaits protocol verification |
| Secondary | mPFC SELF vs WORD | V0Pre, V0Post, week 8 | Week 8 and change; available v909 model covariate-adjusted | Registered; unadjusted rerun required |
| Exploratory/symptom | MADRS | Baseline, week 8 | Week 8 and change | Hypothesis-generating; not efficacy-confirmatory |
| Exploratory/symptom | QIDS-SR16 | Baseline, week 8 | Week 8 and change | Hypothesis-generating |
| Feasibility | Packet-count adherence | Across treatment | Active−placebo average marginal effect | Corrected v729 analysis |
| Biochemical | Serum D-BHB | 0, 20, 40, 80 min at week 8 | Descriptive arm profiles | No prospectively defined exposure target |
| Safety | Adverse events | Through data freeze | Counts by exposed arm | N=26; interim free-text terms |

#### S4. Neuroimaging acquisition, tasks, and processing

MRI used a Siemens 3-T Prisma and 20-channel head coil. BOLD EPI parameters were TR 1500 ms, TE 27 ms, flip angle 72°, matrix 96, field of view 230 mm, 2.4-mm isotropic voxels, 54 slices, and multiband factor 2. MID comprised two runs of 375 volumes; the self-referential adjective task comprised 476 volumes. T1-weighted MPRAGE parameters were TR 1690 ms, TE 2.4 ms, flip angle 8°, 1-mm isotropic resolution, and 256 slices.

AFNI 24.3.10 preprocessing included quintic-interpolated slice timing, rigid-body motion correction, skull stripping, rigid T1-to-EPI registration, 12-parameter affine normalization to MNI space, and 4-mm FWHM smoothing. The pipeline did not include distortion correction, physiological denoising, temporal filtering, or global-signal regression. Volumes were censored at ENORM >0.3 mm or outlier fraction >0.1; a run was discarded when mean ENORM >0.3 mm. First-level AFNI BLOCK models included motion parameters and four Legendre polynomial nuisance terms. Contrasts were MID P5 versus P0 and adjective SELF versus WORD. Bilateral regions were independently defined using Brainnetome v1.0; extracted AFNI-scaled beta coefficients approximate percent signal change.

#### S5. Analysis populations and missing data

Table S4. Population accounting

| **Population/status** | **Overall, n** | **Active, n** | **Placebo, n** | **Definition/note** |
| --- | --- | --- | --- | --- |
| Assessed | 62 | — | — | Eligibility assessment |
| Randomized | 29 | 15 | 14 | Allocation population |
| Never dosed | 3 | 3 | 0 | All three active: claustrophobia |
| Safety/exposed | 26 | 12 | 14 | Received ≥1 dose |
| Exposed but not evaluable | 4 | 2 | 2 | Intervention/imaging, scheduling, or blood-draw limitation |
| Evaluable/primary analysis | 22 | 10 | 12 | Analysis set |

All 22 evaluable participants contributed primary NAcc data at V0Pre, V0Post, and week 8 (66 observations). The secondary mPFC dataset had 61 observations: 22 at V0Pre, 20 at V0Post, and 19 at week 8. Missingness affected two participants per arm across post-dose or week 8 visits. Mixed models used available observations and assumed missing at random; no imputation or pattern-mixture sensitivity analysis was documented. MADRS, QIDS-SR16, and adherence were complete in the evaluable set. D-BHB was available for 21 participants, with one additional missing 80-minute draw in active treatment.

#### S6. Interim-analysis status and decision chronology

The protocol specified an interim analysis once 10 participants entered the per-protocol dataset, with no formal efficacy, futility, safety, or target-engagement boundaries. The governing protocol was version 3.0 (30 October 2023). The present Bayesian analysis includes 22 evaluable participants and was run after the December 2025 data freeze. Accordingly, the report is classified as an exploratory Bayesian interim analysis while preserving the registered status of the outcomes and timepoints. The interim results were not used to determine continuation and were intended for preliminary reports.

#### S7. Bayesian model specification

Table S5. Reproducible model summary and unresolved conflicts

| **Outcome** | **Model** | **Posterior estimand** | **Participants/observations** | **Provenance note** |
| --- | --- | --- | --- | --- |
| Primary NAcc | Gaussian identity LMM: outcome ~ arm × visit + (1\|participant) | Week 8; change | 22/66 | Executable sets iter=4000 including 2000 warm-up; defaults otherwise not printed |
| Secondary mPFC | Same + age + sex + BMI + medication | Week 8; change | 22/61 | Unadjusted main; covariate and certainty diagnosis |
| MADRS | Gaussian identity LMM: score ~ arm × visit + (1\|participant) | Week 8; change | 22/44 | Unadjusted main; covariate and certainty diagnostics |
| QIDS-SR16 | Gaussian identity LMM: score ~ arm × visit + (1\|participant) | Week 8; change | 22/44 | Unadjusted main; covariate diagnostics |
| Adherence | One-inflated beta via constrained zero-one-inflated beta; arm in mean/boundary components | Average marginal effect | 22 | capped primary sensitivity |

Contrasts are active minus placebo. Equal-tailed 95% CrIs are reported for LMMs; adherence output is labeled as 95% highest-density interval in the executable report. No confirmatory posterior decision criterion or multiplicity adjustment was specified. Exploratory covariates were age, sex, BMI, and medication use. Drug-certainty adjustment was post-randomization and is interpreted only as a masking diagnostic.

Priors and sampling conflict across sources. The model-specification memo describes fixed-effect N(0,1), gamma random-intercept variance, exponential residual variance, four chains, and 4000 iterations (2000 warm-up). In the executable report, custom prior arguments are commented out, implying package defaults, and the primary call explicitly sets 4000 iterations; chains and warm-up are not printed. The adherence model prints flat arm coefficients, Student-t(3,0,2.5) intercept priors for mean/precision, and logistic(0,1) for one-inflation.

#### S8. Adherence calculation and sensitivity

Adherence (%) = 100 × (total packets dispensed − total packets returned) / [3 × (V4 date − V0 date) − 2]. The −2 represents two packets administered in clinic at baseline. The interval ranged 52–63 days. All replacements were included in aggregated dispensed and returned counts. One 100.6% value reflected one more packet apparently consumed than expected; reconciliation found no data-entry or denominator error. Because capping was not prespecified, both capped and excluded analyses are reported. Self-reported MARS/MSAQ and MEMS-cap adherence metrics were absent from the interim dataset.

#### S9. Biochemical exposure

D-BHB was measured at pre-dose and 20, 40, and 80 minutes at week 8. LLOQ was 0.05 mmol/L and values below detectable limit were imputed as 0.05/√2. No target concentration was prospectively defined, so profiles are interpreted as biochemical separation, not adequacy. Arm patterns revealed which A/B label contained active supplement and therefore created de facto unmasking.

#### S10. Safety methods

The safety population included all 26 exposed participants. Events were captured through the December 19, 2025 freeze; the latest recorded event occurred at week 10, but the exact last safety follow-up date is unknown. Severity used protocol-defined categories; all four events were mild. Relationship was assessed by the investigator/study physician; no event was possibly, probably, or definitely related. Terms remained free-text and were not MedDRA coded.

### Supplementary Results

#### S11. Bayesian results and sensitivity analyses

Table S6. Main and selected sensitivity estimates

| **Outcome** | **Analysis** | **Estimand** | **Median (95% CrI/HDI)** |
| --- | --- | --- | --- |
| NAcc primary | Unadjusted | Week 8 | 0.02 (−0.12, 0.17) |
| NAcc primary | Unadjusted | Change | 0.00 (−0.17, 0.18) |
| mPFC secondary | Unadjusted | Week 8 | 0.00 (−0.18, 0.18) |
| mPFC secondary | Unadjusted | Change | 0.08 (−0.15, 0.33) |
| MADRS | Unadjusted | Week 8 | −7.28 (−12.00, −2.47) |
| MADRS | Unadjusted | Change | −7.12 (−12.80, −1.52) |
| MADRS | Covariate adjusted | Week 8 | −7.87 (−12.80, −3.14) |
| MADRS | Covariate adjusted | Change | −7.15 (−12.80, −1.68) |
| MADRS | Drug-certainty diagnostic | Week 8 | −5.49 (−13.50, 2.34) |
| MADRS | Drug-certainty diagnostic | Change | −3.38 (−9.47, 2.86) |
| MADRS | Imaging-complete subset | Week 8 | −5.75 (−11.20, −0.31) |
| MADRS | Imaging-complete subset | Change | −6.44 (−12.80, 0.29) |
| QIDS-SR16 | Unadjusted | Week 8 | −1.95 (−5.13, 1.15) |
| QIDS-SR16 | Unadjusted | Change | −1.88 (−4.88, 1.13) |
| QIDS-SR16 | Covariate adjusted | Week 8 | −1.82 (−5.30, 1.64) |
| QIDS-SR16 | Covariate adjusted | Change | −1.90 (−4.79, 1.05) |
| Adherence | 100.6% capped | AME | 0.0117 (−0.0706, 0.0941) |
| Adherence | 100.6% excluded | AME | 0.0013 (−0.0863, 0.0837) |

Active minus placebo. AME, average marginal effect. The 'imaging-complete subset' is not a complete-case symptom analysis because symptom data were complete; it restricts the symptom model to participants with complete imaging. Drug certainty is post-randomization and changes the estimand.

#### S12. Model diagnostics

Table S7. Reported diagnostics

| **Model** | **Maximum R-hat** | **Minimum bulk ESS** | **Minimum tail ESS** | **Divergences / treedepth** |
| --- | --- | --- | --- | --- |
| Primary NAcc | 1.002 | 1442.6 | 2042.8 | 0 / 0 |
| Secondary mPFC (adjusted) | 1.003 | 3395.8 | 3773.8 | 0 / 0 |
| MADRS | 1.004 | 1571.1 | 2929.3 | 0 / 0 |
| QIDS-SR16 | 1.001 | 1917.2 | 2770.6 | 0 / 0 |
| Adherence, capped/truncated | 1.001 | 3530.2 | 3920.6 | 0 / 0 |
| Adherence, excluded | Not independently verified | — | — | — |

The executable code calls the capped/truncated adherence object for both excluded and truncated diagnostic rows and posterior predictive checks; excluded-model diagnostics are therefore not independently verified. ESS, effective sample size.

#### S13. D-BHB and adherence results

Table S8. Week 8 D-BHB: median (range) [quantifiable/n]

| **Time** | **Active** | **Placebo** |
| --- | --- | --- |
| Pre-dose (0 min) | <0.05 (<0.05–0.09) [3/10] | <0.05 (<0.05–0.27) [5/11] |
| 20 min | 0.15 (0.05–0.28) [10/10] | <0.05 (<0.05–0.25) [5/11] |
| 40 min | 0.16 (0.08–0.28) [10/10] | 0.05 (<0.05–0.20) [6/11] |
| 80 min | 0.09 (<0.05–0.16) [7/9] | <0.05 (<0.05–0.08) [5/11] |
| Peak post-dose | 0.18 (0.08–0.28) [10/10] | <0.05 (<0.05–0.25) [6/11] |

LLOQ=0.05 mmol/L. Below-LLOQ values are displayed as <0.05 without substitution. One placebo participant had no D-BHB data; one active participant lacked the 80-min draw.

Table S9. Packet-count adherence

| **Group** | **n** | **Mean (SD), as computed** | **Median** | **Range** | **Mean (SD), capped** |
| --- | --- | --- | --- | --- | --- |
| Active | 10 | 90.1 (13.0) | 97.2 | 65.6–100.6 | 90.1 (13.0) |
| Placebo | 12 | 91.9 (7.2) | 92.8 | 78.9–100.0 | 91.9 (7.2) |
| Overall | 22 | 91.1 (10.0) | 94.2 | 65.6–100.6 | 91.0 (10.0) |

#### S14. Imaging quality control

QC was run while masked. The only clearly notable arm imbalances occurred at V0Pre, before exposure: censored TR fractions were higher in active treatment for MID and the adjective task, and maximum adjective-task motion was higher. Mean motion remained below the 0.3-mm run-exclusion threshold at every visit. P values are descriptive and multiplicity-unadjusted.

Table S10. MID task QC

| **Metric** | **Visit** | **Active mean (SD)** | **Placebo mean (SD)** | **t-test P** | **Wilcoxon P** |
| --- | --- | --- | --- | --- | --- |
| Mean motion, mm | V0Pre | 0.138 (0.055) | 0.116 (0.028) | 0.278 | 0.277 |
|  | V0Post | 0.154 (0.062) | 0.132 (0.039) | 0.351 | 0.373 |
|  | V4 | 0.166 (0.074) | 0.138 (0.039) | 0.299 | 0.621 |
| Maximum motion, mm | V0Pre | 4.01 (1.60) | 3.81 (2.03) | 0.803 | 0.817 |
|  | V0Post | 3.98 (2.28) | 4.47 (1.80) | 0.586 | 0.307 |
|  | V4 | 4.68 (2.77) | 5.07 (3.73) | 0.782 | 0.974 |
| TRs censored, % | V0Pre | 13.9 (11.5) | 5.4 (5.8) | 0.055 | 0.025 |
|  | V0Post | 19.6 (17.8) | 9.3 (7.5) | 0.115 | 0.147 |
|  | V4 | 21.3 (23.8) | 9.0 (8.8) | 0.152 | 0.129 |
| Response rate, % | V0Pre | 54.9 (10.5) | 60.5 (10.7) | 0.233 | 0.129 |
|  | V0Post | 58.4 (11.3) | 55.6 (11.0) | 0.565 | 0.448 |
|  | V4 | 57.3 (8.4) | 54.7 (16.5) | 0.638 | 0.791 |

Table S11. Self-referential adjective-task QC

| **Metric** | **Visit** | **Active mean (SD)** | **Placebo mean (SD)** | **t-test P** | **Wilcoxon P** |
| --- | --- | --- | --- | --- | --- |
| Mean motion, mm | V0Pre | 0.186 (0.049) | 0.153 (0.040) | 0.107 | 0.138 |
|  | V0Post | 0.186 (0.079) | 0.194 (0.082) | 0.824 | 0.621 |
|  | V4 | 0.192 (0.094) | 0.191 (0.072) | 0.973 | 0.410 |
| Maximum motion, mm | V0Pre | 5.04 (2.64) | 2.68 (1.02) | 0.022 | 0.060 |
|  | V0Post | 3.13 (2.15) | 3.17 (2.18) | 0.970 | 0.668 |
|  | V4 | 3.31 (1.96) | 3.76 (2.77) | 0.661 | 0.767 |
| TRs censored, % | V0Pre | 26.2 (14.3) | 11.4 (9.0) | 0.013 | 0.013 |
|  | V0Post | 25.2 (25.2) | 19.3 (14.9) | 0.523 | 0.974 |
|  | V4 | 24.7 (27.4) | 21.7 (19.5) | 0.781 | 0.717 |
| Response rate, % | V0Pre | 90.9 (23.7) | 95.0 (6.2) | 0.613 | 0.219 |
|  | V0Post | 94.2 (9.7) | 86.0 (19.1) | 0.215 | 0.452 |
|  | V4 | 90.2 (17.0) | 86.6 (17.7) | 0.636 | 0.548 |

#### S15. Adverse events

Table S12. Adverse events by arm (safety population)

| **Category** | **Active (n=12)** | **Placebo (n=14)** | **Total (N=26)** |
| --- | --- | --- | --- |
| Participants with any AE | 1 (8.3) | 2 (14.3) | 3 (11.5) |
| Total AEs, events | 1 | 3 | 4 |
| Treatment-related AEs | 0 | 0 | 0 |
| Serious AEs | 0 | 0 | 0 |
| Severe AEs | 0 | 0 | 0 |
| Dose interruption/discontinuation/withdrawal | 0 | 0 | 0 |
| Deaths | 0 | 0 | 0 |
| Suicidal ideation/behavior | 1 (8.3) | 2 (14.3) | 3 (11.5) |
| Constitutional event | 0 | 1 (7.1) | 1 (3.8) |

Table S13. Event-level adverse-event listing

| **Participant** | **Arm** | **Visit** | **Event summary** | **Severity** | **Relationship** | **Outcome** |
| --- | --- | --- | --- | --- | --- | --- |
| BW881 | Placebo | Week 8 | Feeling unwell during MRI practice; low glucose/ear congestion; resolved with food/rest | Mild | Unlikely | Resolved |
| BW881 | Placebo | Week 10 | Transient passive wish not to awaken; denied to physician; no intent/plan/behavior | Mild | Not related | Resolved |
| BZ190 | Placebo | Week 2 | Passive death wishes with sadness; no intent or plan | Mild | Not related | Resolved |
| BD704 | Active | Week 8 | Brief controllable self-harm urges; physician assessed low risk; no intent/plan/behavior | Mild | Not related | Ongoing at report |

Identifiers are study codes from the supplied de-identified listing. No event was serious. Terms were not MedDRA coded.

#### S16. Analysis deviations and transparency summary

Table S14. Material deviations or unresolved status

| **Domain** | **Planned/earlier state** | **Actual/current state** | **Treatment in package** |
| --- | --- | --- | --- |
| Statistical framework | Original plan appears non-Bayesian; protocol absent | Bayesian LMM/OIB | Classified exploratory; protocol/code required |
| Interim sample | Approx. 10 per-protocol participants described | 22 evaluable (10/12) | Expanded interim dataset disclosed |
| Population | Protocol definition not directly verified | Evaluable/per-protocol subset | 29/26/22 populations separated |
| Secondary model | Unadjusted expected from model memo | Only covariate-adjusted v909 output | Labeled adjusted; rerun required |
| Priors/sampling | Conflicting memo and executable report | Package defaults/custom settings unclear | No invented resolution; author query |
| Adherence | Earlier negative arm difference | Corrected near-zero positive AME | Version 909 used |
| BHB censoring | v729 substitutes LLOQ/√2 | Memo tabulates <LLOQ | Methods disclose both |
| Masking | Intended masking | BHB pattern caused de facto unmasking | Timing and dissemination queried |
| Safety denominator | Earlier narrative incomplete | 26 exposed | Complete interim AE table included |

Table S15. Transparency items requiring author completion

| **Area** | **Required completion** |
| --- | --- |
| Protocol/SAP | Protocol v3.0, amendments, and exact section 9.4.6 language |
| Analysis provenance | Decision date/rationale; who saw what data; exact code, priors, chains, iterations, warm-up |
| Registration | Direct registry history; timing of first enrollment vs public registration |
| Oversight | IRB name/number/date; protocol deviations; monitoring |
| Masking | Unmasked personnel and dates; access to A/B key; result recipients |
| Safety | Exact last follow-up; MedDRA version/coding; update through submission |
| Adherence | MARS/MSAQ and MEMS reconciliation |
| Administration | Funding, company role, product/material support, any equity-based support and recipients, conflicts, author contributions |
| Open science | Data/code repository, terms, release timing, model objects, session information |

#### S17. Formulation composition and analytical testing

The active study intervention was Brain Energy Complex, a multicomponent metabolic formulation developed and supplied by KetoSwiss. Each stick pack was formulated to contain D-β-hydroxybutyrate (D-BHB) as calcium and magnesium salts together with micronutrients and metabolic cofactors. The target formulation and quantities per serving used for the study are shown in Table S16.

Independent analytical testing of the study product was performed by Dyad Labs (Mérieux NutriSciences) on study lot 23214A. Further results for the tested sample are shown separately in Table S17. Minor differences between analytically measured values and target formulation values are expected as a result of normal manufacturing and analytical variability. The analyzed sample had a measured serving weight of 9.2 g, compared with the nominal approximately 9.25-g stick-pack weight described in the study protocol.

Table S16. Reported composition of the supplied Brain Energy Complex sample

| **Analyte or reported form** | **Unit** | **Result per 9.2-g serving** | **Calculated in three servings** |
| --- | --- | --- | --- |
| Beta carotene | µg | 81.3 | 243.9 |
| Carnitine tartrate (calculated) | mg | 183 | 549 |
| Citicoline sodium | mg | 12.2 | 36.6 |
| Boron (B) | µg | 340 | 1020 |
| Calcium (Ca) | mg | 417 | 1251 |
| Chromium (Cr) | µg | 31.6 | 94.8 |
| Magnesium (Mg) | mg | 275 | 825 |
| Manganese (Mn) | mg | 0.968 | 2.904 |
| Molybdenum (Mo) | µg | 13.9 | 41.7 |
| Potassium (K) | mg | 216 | 648 |
| Selenium (Se) | µg | 27.6 | 82.8 |
| Sodium (Na) | mg | 130† | 390 |
| Zinc (Zn) | mg | 5.72 | 17.16 |
| Total tocopherols | mg | 13.8† | 41.4 |
| Ascorbic acid | mg | 60.6 | 181.8 |
| Leucine | mg | 49.4† | 148.2 |
| Lysine | mg | 46.5† | 139.5 |
| Lysine HCl | mg | 58.1 | 174.3 |
| Taurine | mg | 165 | 495 |
| beta-Hydroxybutyrate | mg | 4910 | 14730 |
| Carnitine HCl | mg | 153 | 459 |
| Carnitine | mg | 125 | 375 |
| Methylcobalamin | µg | 83.7 | 251.1 |
| Niacin | mg | 2.71 | 8.13 |
| Niacinamide | mg NE | 33.5 | 100.5 |
| Pyridoxal-5-phosphate | mg | 2.48† | 7.44 |
| Thiamine HCl | mg | 3.54 | 10.62 |
| alpha-Tocopherol acetate | mg | 6.03 | 18.09 |
| Menaquinone 7 (K2) | µg | 10.6† | 31.8 |
| Phylloquinone (K1) | µg | 12.6 | 37.8 |
| Retinyl palmitate | µg | 146 | 438 |
| Ca 5-methyltetrahydrofolic acid | µg DFE | 79.2† | 237.6 |
| 5-Methyltetrahydrofolic acid | µg DFE | 73.1† | 219.3 |
| Biotin | µg | 32.1 | 96.3 |
| Calcium pantothenate | mg | 12.5 | 37.5 |
| Cholecalciferol (D3) | µg | 23.5† | 70.5 |
| Iodine (I) | µg | 26.6 | 79.8 |
| Riboflavin-5-phosphate | mg | 15.6 | 46.8 |
| Ubiquinol | mg | 49.6† | 148.8 |

**Notes to Table S16**

Values are transcribed from the certificate. The three-serving column is calculated as three times each reported per-serving result; it is not a separate measurement or a confirmed participant dose. No adjustment from the certificate’s 9.2-g serving to the manuscript’s approximately 9.25-g packet was made.

† Reported result is numerically below the certificate’s stated minimum. These flags describe the comparison in the supplied report and do not constitute an independent product-release or clinical-safety determination. Specifications are not measured quantities or intended ingredient doses.

NE, niacin equivalents; DFE, dietary folate equivalents; HCl, hydrochloride. Original units are retained. Elemental minerals may include mineral counterions. Entries for carnitine, lysine, folate, and tocopherol forms or equivalents may overlap and must not be summed to infer total dose or ingredient count. Carnitine tartrate is identified by the report as a chromatography calculation. The source misspelling “Ubiquionol” is rendered as ubiquinol, consistent with its test-method label.

Table S17. Other analytical results in the supplied certificate

| **Test** | **Unit** | **Reported result** |
| --- | --- | --- |
| Yeasts and molds | CFU/g | <10 |
| Aerobic plate count | CFU/g | <10 |
| Coliforms | CFU/g | <10 |
| Arsenic (As) | ppm | 0.0226 |
| Cadmium (Cd) | ppm | 0.0101 |
| Lead (Pb) | ppm | 0.0539 |
| Mercury (Hg) | ppm | <0.00905 |
| Loss on drying | % w/w | 2.18 |

CFU/g, colony-forming units per gram; ppm, parts per million; w/w, weight/weight. Concentrations and microbial counts are not multiplied by the daily serving frequency. The complete source certificate accompanies the revised manuscript package. Its page-one header lists certificate ID 1287825-1, whereas the page footers list 1287825-2; the shared laboratory sample ID above identifies the report.

### Supplementary Figures

Figure S1. Biochemical exposure and packet-count adherence

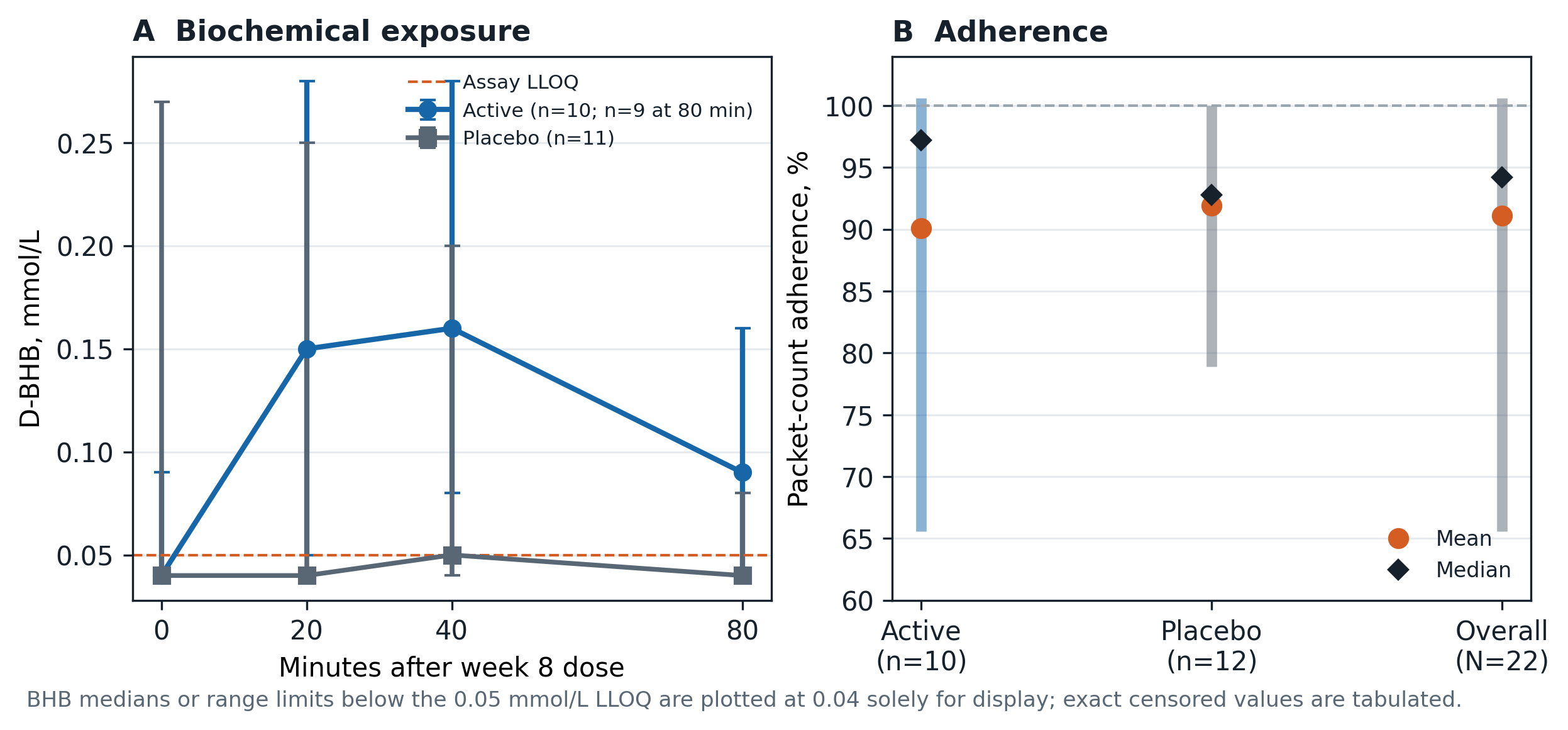

Panel A displays median and range D-BHB. Below-LLOQ values are assigned a plotting position of 0.04 mmol/L only for visualization; exact censored values appear in Table S8. Panel B displays means, medians, and ranges. The 100.6% value was verified and retained descriptively.

Figure S2. Adherence sensitivity analyses

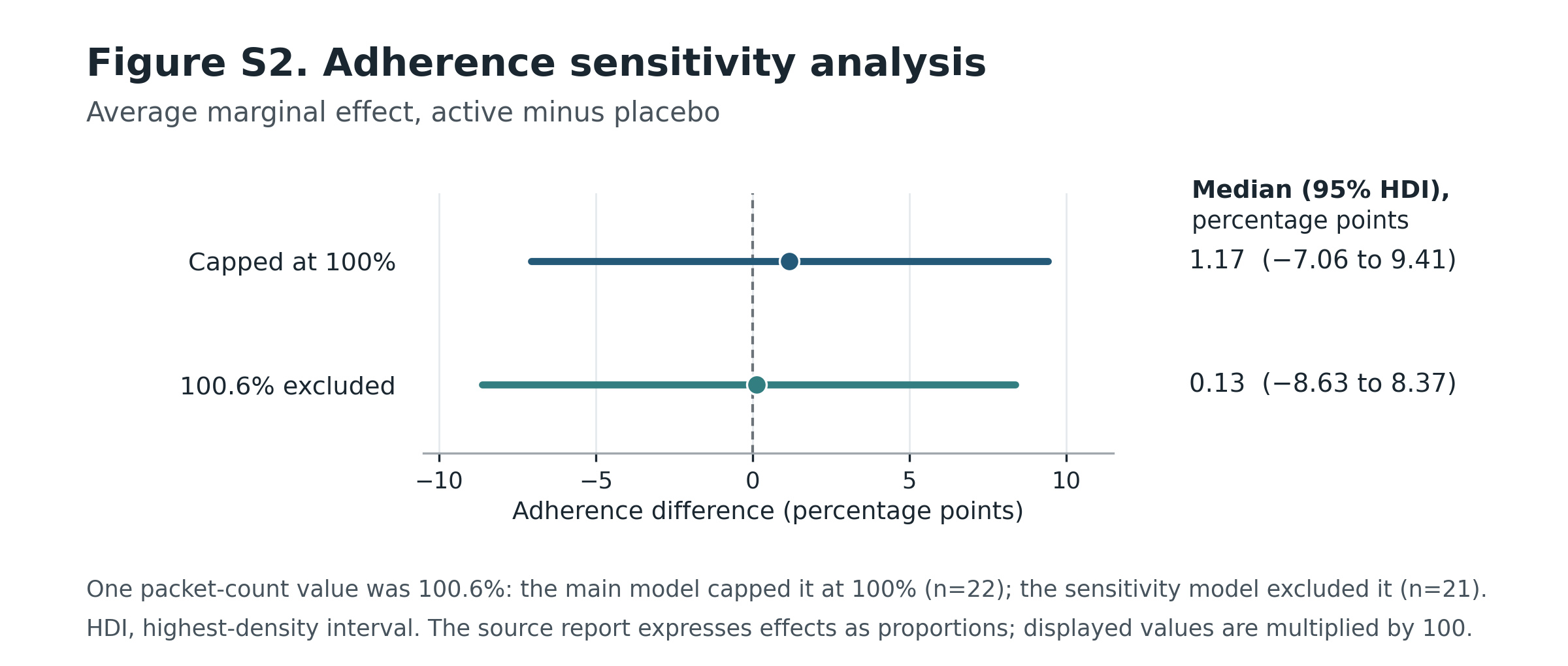

Posterior medians and 95% intervals for active minus placebo average marginal effects from one-inflated beta models. The executable code contains reuse errors in adjusted/excluded diagnostics; only these unadjusted reported contrasts are reproduced.

Figure S3. Posterior predictive checks

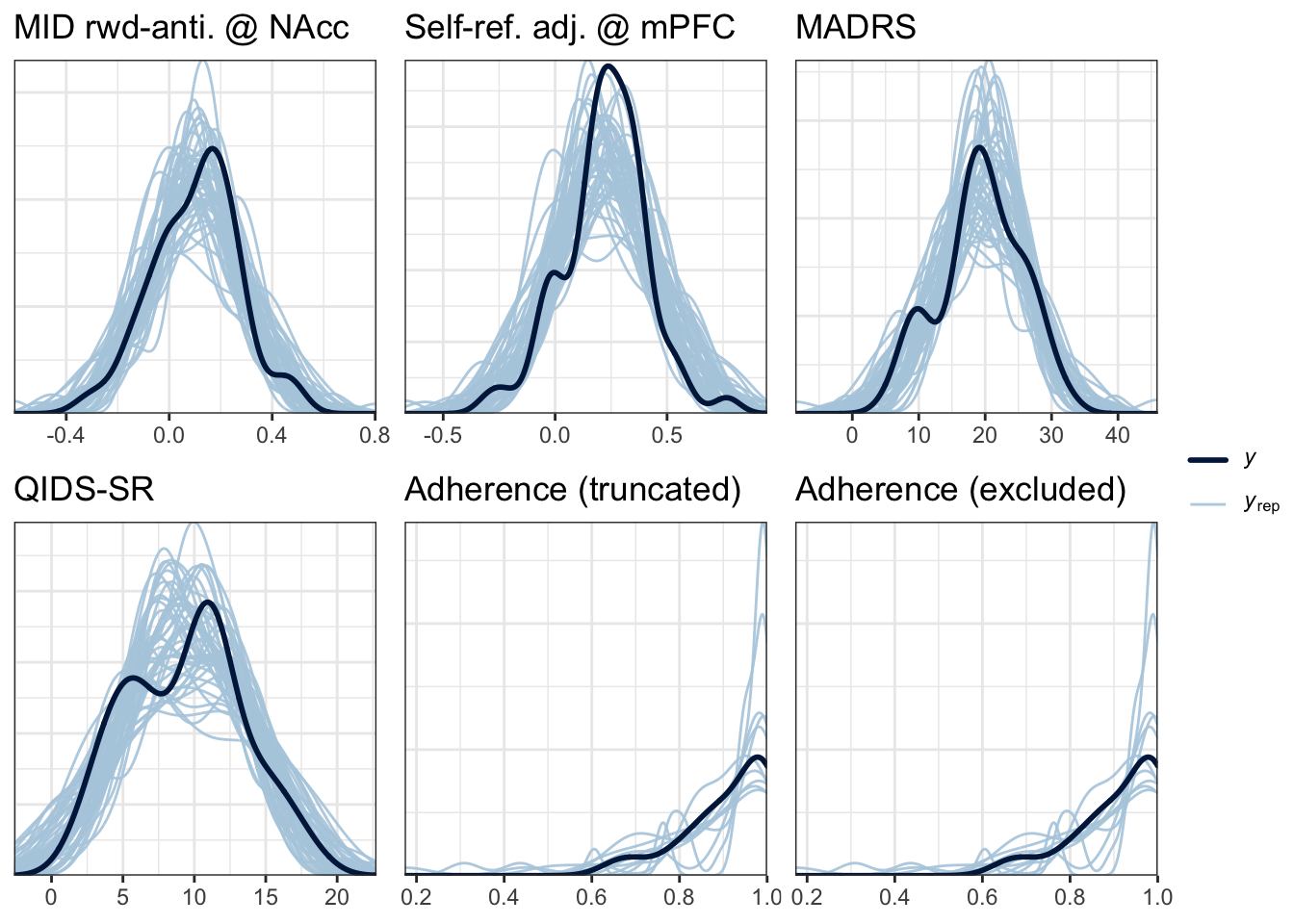

Observed density (dark) and replicated draws (light) for the primary, adjusted secondary, MADRS, QIDS-SR16, and capped/truncated adherence models. The excluded-adherence panel is omitted because the code reused the truncated model, so it was not independently verified.
